# Incidence and risk factors associated with acquired syphilis in HIV pre-exposure prophylaxis users

**DOI:** 10.1101/2024.04.25.24306355

**Authors:** Nathália Lima Pedrosa, Patrícia Matias Pinheiro, Fernando Wagner Brito Hortêncio Filho, Wildo Navegantes de Araujo

**Affiliations:** Tropical Medicine Center, University of Brasília, Brasília, Federal District, Brazil; Walter Cantídio University Hospital, Federal University of Ceará, Fortaleza, Ceará, Brazil; Federal Institute of Education, Science, and Technology of Brasília, Brasília, Federal District, Brazil; UnB Faculty Ceilândia, University of Brasilia, Brasília, Federal District, Brazil; National Institute for Science and Technology for Health Technology Assessment, Porto Alegre, RS, Brazil

**Keywords:** Syphilis, HIV, PrEP, Cohort, Brazil

## Abstract

**Background:** Acquired syphilis continues to affect millions of people around the world. It is crucial to study it in the context of HIV Pre-Exposure Prophylaxis (PrEP) to achieve the goals set out in the 2030 Agenda since the literature suggests increased risk behaviors for sexually transmitted infections. This study aimed to investigate the incidence and factors associated with acquired syphilis among PrEP users.

**Materials and methods:** This retrospective cohort included data on PrEP users from all over Brazil from 2018 to 2020, obtained from the national antiretroviral logistics system. We calculated the proportion of syphilis before PrEP, the incidence during the user’s follow-up, reinfections, and their possible associated factors. We conducted descriptive, bivariate, and multivariate analysis, estimating the crude Relative Risk, adjusted Odds Ratio (aOR), and their respective confidence intervals (95%CI).

**Results:** Most of the 34,000 individuals who started PrEP were male (89.0%), white (53.7%), self-identified as male (85.2%), homosexual, gay, or lesbian (72.2%), and had 12 schooling years or more (67.8%). Of these, 8.3% had syphilis in the six months before starting PrEP, and 4% had it in the first 30 days of using the prophylaxis. In the 19,820 individuals effectively monitored, the incidence of acquired syphilis was 19.1 cases per 100 person-years, and 1.9% of users had reinfection. The multivariate analysis identified female gender (aOR 0.3; 95%CI 0.2-0.5), being white or Black (aOR 0.9; 95%CI 0.7-0.9 and aOR 0.7; 95%CI 0.7-0.99, respectively) as protective factors for syphilis. Being homosexual, gay, lesbian (aOR 2.7; 95%CI 2.0-3.7) or having a history of syphilis in the six months before PrEP (aOR 2.2; 95%CI 1.9-2.5) were risk factors for syphilis during PrEP use. Behaviors related to the risk of syphilis included accepting something in exchange for sex (aOR 1.6; 95%CI 1.3-1.9), irregular condom use (use in less than half of sexual intercourse sessions; aOR 1.7; 95%CI 1.53-2.1) and recreational drug use (poppers; aOR 1.5; 95%CI 1.53-2.1).

**Conclusion:** Syphilis in the context of PrEP has high rates and is associated with sociodemographic and behavioral factors. We recommend additional studies targeting prevention in this population to curb these figures.

## INTRODUCTION

Acquired syphilis is the most common sexually transmitted infection (STI) worldwide. Although it has been recognized since the 15^th^ century, it continues to represent a contemporary challenge, affecting millions of people, with an annual estimate of 7.1 million (3.8-10.3 million) new infections between 2016 and 2020 worldwide [1–3].

Syphilis is caused by the spirochete bacterium *Treponema pallidum*. It can result in severe health complications and genital symptoms, including complications during pregnancy, infertility, increased risk of infection by the human immunodeficiency virus (HIV), and psychological impacts if not appropriately treated. The incidence is significantly higher in men who have sex with men (MSM) and in people living with HIV (PLHIV). Appropriate management involves staging the disease, treatment with benzathine penicillin for the individual and their partners, screening in at-risk populations, and notification to health authorities to support control and prevention [2–5].

In response to this enduring public health issue, syphilis has been incorporated into the Sustainable Development Goals of Agenda 2030, launched by the United Nations (UN), and the Strategic Guidelines for the Elimination of Sexually Transmitted Infections by the World Health Organization (WHO), which aim to reduce the incidence of *T. pallidum* infections by 90% [1].

Besides this setting, Pre-Exposure Prophylaxis (PrEP) has emerged as a crucial strategy in the fight against HIV. Launched in 2012 and widely disseminated globally, the provenly highly effective PrEP involves the use of oral antiretrovirals (ARVs) (in daily doses or on-demand) to reduce the risk of infection by the virus [6]. While PrEP has reduced HIV worldwide, the low use of condoms in individuals using prophylaxis and other “risk compensations” [7] may pose additional challenges in reducing syphilis and achieving the goals set for the 2030 Agenda.

Despite the extensive research on PrEP and STIs, there is a dearth of studies analyzing acquired syphilis, its recurrence among PrEP users, and the factors associated with its incidence. These findings could provide critical evidence to strengthen strategies to combat syphilis and help achieve the targets set by the WHO. Therefore, this study aimed to analyze the incidence of acquired syphilis in the population of PrEP users and identify the factors associated with this incidence.

## MATERIALS AND METHODS

### Design

This retrospective cohort employed secondary longitudinal data from the Brazilian Ministry of Health’s Medicines Logistics Control System (SICLOM), which is used nationwide to manage the distribution of antiretroviral drugs. This system keeps records of ARV users, both for treatment and prophylaxis, along with clinical and laboratory information on the follow-up of these users [8].

The databases extracted from SICLOM, containing information from standardized appointments with PrEP users, were provided by the Ministry of Health after the study was approved by the Ethics Committee of the Faculty of Medicine of the University of Brasília (CAAE: 07448818.0.1001.5558). The data was made available to researchers on March 7, 2021 and were anonymized by removing names, dates of birth, and addresses before being made available to the researchers and replaced by untraceable numerical codes for each study participant in order to guarantee information confidentiality and the protection of PrEP users.

During the period in which the database was available (2018-2020) in Brazil, the guidelines standardized care for PrEP users, recommending an initial appointment to collect sociodemographic and behavioral risk data and clinical and laboratory investigation for HIV and other sexually transmitted infections (STIs), using specific forms (Supplement 1). After the first return in 30 days, PrEP users should be followed up every three months for clinical and laboratory follow-up [9]. During the study period, the Unified Health System performed evaluation and follow-up appointments, laboratory tests, and the dispensing of antiretroviral drugs for PrEP at no cost to the user [10].

A database was compiled for each form used between 2018 and 2020. The process of qualifying the database of PrEP users is described in Supplement 2. It allowed for following the PrEP user’s trajectory from admission to the last available follow-up appointment.

### Study population

Considering the target study population as PrEP users, we selected individuals who met the eligibility criteria in the national policy in force at the time [9] for prophylaxis in Brazil, seen between January 2018 and December 2020 and who had at least one follow-up visit. Afterward, users for whom it was impossible to obtain information from any subsequent forms were excluded through a deterministic relationship between the different databases created from the records derived from the completed forms (Supplement 2).

In Brazil, the PrEP eligibility criteria include sexually active people with increased risk for HIV, such as sexual practices with penetration without the use of condoms, multiple casual sexual partners, diversity and frequency of sexual partnerships, history of STIs, repeated search for HIV Post-Exposure Prophylaxis (PEP), sexual relationships in exchange for money or material goods, and Chemsex (sexual practice under the influence of psychoactive drugs).

While not exclusive, serodiscordant couples and primary populations (gay men, men who have sex with men (MSM), transgender people, and sex workers) should be given preference in the provision of PrEP by the Unified Health System (SUS) [9].

### Measures

Data collected consisted of sociodemographic characteristics (genitalia at birth, self-reported ethnicity/skin color, gender identity, sexual orientation, age, and schooling years), behavioral characteristics in the last three months (sex in exchange for money/goods, condom use frequency, alcohol consumption, use of injectable drugs and other psychoactive drugs, number and type of sexual partnerships), and health characteristics (history of STIs in the last six months and presence of active syphilis during follow-up). Self-reported ethnicity/skin color is used per the Brazilian Institute of Geography and Statistics (IBGE). We arbitrated the information referring to the last appointment for follow-up behavioral data.

In Brazil, the diagnosis of syphilis is guided by clinical evaluation and direct (when with symptoms) or immunological (regardless of symptoms) tests. Concerning the latter, which are more common in routine services, guidelines recommend starting the investigation with a treponemal test (preferably a rapid test) followed by a non-treponemal test (VDRL or RPR) when the first test is positive [11]. The history of acquired syphilis was assessed in the six months before the first appointment through the report of diagnosis or the syphilis serological screening on admission.

In this cohort, the duration of follow-up for PrEP users was calculated as the time elapsed between the date of the first visit and the date of the last follow-up visit, regardless of adherence. Moreover, users who did not attend at least one follow-up appointment were considered losses in the cohort and compared with those who did follow up regarding schooling, ethnicity/skin color, and sexual orientation to check for differences between the groups.

The incidence of acquired syphilis during PrEP use was measured by the number of “active syphilis” events identified by health professionals after three months of prophylaxis use, obtained from the data on the Monitoring Form, following the method described by Ong [12]. The outcome could also be observed on more than one occasion during the user’s follow-up, and this event was defined as a syphilis reinfection outcome during PrEP follow-up and calculated as another incident event. We could, thus, measure the proportion of users who acquired syphilis at least twice or more during PrEP.

### Statistics

We presented the continuous variables (age and number of sexual partners) using measures of central tendency (mean and median) and dispersion (standard deviation and interquartile range), and the categorical variables were described using measures of absolute and relative frequency in each category. The individual’s time in the cohort was calculated as the difference between the date of registration in the system (when PrEP started) and the date of the last appointment recorded in the system. The user follow-up period and syphilis incidence were calculated per 100 person-years.

Sociodemographic and behavioral variables were compared between individuals diagnosed with syphilis and those who were not diagnosed during follow-up. Subsequently, the bivariate association between the outcome and the covariates was verified using the chi-square or Fisher’s exact test for categorical variables and the Wilcoxon test for numerical variables, considering the difference statistically significant if p < 0.05. Furthermore, we calculated the Relative Risk (RR) and the 95% Confidence Interval (95% CI) to evaluate the possible factors associated with syphilis infection during PrEP use.

We then carried out a logistic regression analysis using the backward-stepwise method to control for confounders. In this regression, we considered active syphilis during PrEP (yes or no) as the dependent variable and the sociodemographic, behavioral, and sexual health variables whose bivariate analysis showed a p-value < 0.2 as independent variables. We assessed multicollinearity using Variance Inflation Factors (VIF) and identified the best model using the Akaike Information Criterion (AIC) and pseudo R². Considering the low risk of the outcome in this study, we estimated the Relative Risk adjusted by the adjusted odds ratio (aOR) and its Confidence Intervals (95% CI) to assess the effect of the adjusted covariates on the outcome.

The above analyses were performed using the R® 4.3.3 software using the libraries tidyverse, janitor, stringr, rio, here, purrr, gtsummary, broom, lmtest, parameters, see, readxl, plyr, with code provided as supplementary material (Supplement 2).

## RESULTS

After linking the databases of PrEP consultation forms, we obtained 34,000 records of people who started using PrEP and 19,820 users who followed up and were part of the cohort. The base population of this cohort reflects PrEP users in Brazil between 2018 and 2020, as shown in Table 1. Most PrEP users were male (89.0%), white (53.7%), self-identified as male (85.2%), were homosexuals, gays, or lesbians (72.2%), and had 12 years or more of schooling (67.8%) (Table 1).

**Table 1.** Demographic characteristics of PrEP¹ users for HIV in Brazil (2018-2020).

| **Variables (n)** | **n (%)** |
| --- | --- |
| **Sex at birth(34,000)** |  |
| Male | 30,208 (89.0) |
| Female | 3,771 (11.0) |
| Hermaphrodite | 21 (<0.1) |
| **Self-reported ethnicity/skin color (33,990)** |  |
| White | 18,252 (53.7) |
| Pardo (Brown, i.e., mixed race) | 10,945 (32.2) |
| Black | 4,290 (12.6) |
| Yellow (i.e., Asian Brazilian) | 359 (1.0) |
| Indigenous | 144 (0.4) |
| **Gender identity (34,000)** |  |
| Man | 28,956 (85.2) |
| Transgender man | 103 (0.3) |
| Woman | 3,970 (12) |
| Transgender woman | 790 (2.3) |
| Transvestite | 181 (0.5) |
| **Sexual Orientation (34,000)** |  |
| Bisexual | 3,351 (9.9) |
| Heterosexual | 6,092 (17.9) |
| Homosexual, gay, or lesbian | 24,557 (72.2) |
| **Schooling level<sup>2</sup> (33,991)** |  |
| 1-3 years | 193 (0.6) |
| 4-7 years | 1,616 (4.8) |
| 8-11 years | 9,071 (26.7) |
| 12 years and over | 23,040 (67.8) |
| None, no formal education | 71 (0.2) |

<sup>1</sup> PrEP = Pre Exposure Prophylaxis; <sup>2</sup> Schooling level: school complete years

In total, 19,820 people were effectively followed up, with a median follow-up of seven months (IQR=7-20 months). We observed a loss-to-follow-up rate of 41.7% among those who started PrEP. Similar to the initial cohort, most were men (86%), homosexuals, gays, or lesbians (67%), white (51%), and had 12 years or more of schooling (61%), with p < 0.001.

Table 2 shows an overview of acquired syphilis among PrEP users. During the initial appointment, 8.3% of individuals reported a history of acquired syphilis, and 4.0% of those who returned for follow-up after 30 days were diagnosed with active syphilis.

**Table 2.** Characterization of acquired syphilis before and during follow-up in HIV PrEP users, Brazil, 2018-2020.

| Variables | n (%) |
| --- | --- |
| **Syphilis within 6 months before the initial PrEP appointment <sup>1</sup>(34,000\*)** | 2,806 (8.3) |
| **Active syphilis on return 30 days after initial appointment (19,440\*\*)** |  |
| Yes | 775 (4.0) |
| No | 12,174 (62.6) |
| Not performed/unavailable | 6,491 (33.4) |
| **Active syphilis during HIV PrEP (19,820\*\*)** |  |
| Yes | 1,962 (9.8) |
| No | 17,858 (90.1) |
| **Active syphilis per follow-up appointment (79,753\*\*\*)** |  |
| Yes | 2,525 (3.2) |
| No | 46,222 (58.0) |
| Not performed/unavailable | 30,949 (38.8) |
| **Number of active syphilis episodes per user (19,820\*\*)** |  |
| 0 | 17,858 (90.1) |
| 1 | 1,590 (8) |
| 2 | 290 (1.5) |
| 3 | 65 (0.3) |
| 4 | 15 (<0.1) |
| 5 | 2 (<0.1) |

\* Study base population; \*\* Cohort population; \*\*\* Total follow-up appointments made.<sup>1</sup> PrEP = Pre Exposure Prophylaxis.

In this cohort, 1,962 individuals (9.8%) received at least one diagnosis of active syphilis during follow-up, and 2% of users had a recurring infection, i.e., more than one episode of syphilis during follow-up. In total, 2,525 diagnoses of active syphilis were recorded, resulting in an incidence of 19.1 cases of acquired syphilis per 100 person-years (95% CI 18.4-19.9). There was a high proportion of missed syphilis tests in the 30-day return period (33.4%) and the total number of follow-up appointments (38.8%).

To analyze the risk factors associated with acquired syphilis, we compared individual and behavioral characteristics between people with at least one diagnosis of acquired syphilis during follow-up and PrEP users who did not receive such a diagnosis during the period (see Table 3).

**Table 3.** Characteristics of PrEP users and factors associated with syphilis during follow-up, Brazil, 2018-2020.

| Variable (n) | Syphilis episode during PrEP |  | RR* | 95% CI | p-value *** | aOR# | 95%CI## | p-value *** |
| --- | --- | --- | --- | --- | --- | --- | --- | --- |
|  | Yes n (%)<br>(n=1,962) | No n (%)<br>(n=17,858) |  |  |  |  |  |  |
| **Age (19,811)** | 32 ± 8 (31)** | 33 ± 9 (31)** |  |  | 0.366 |  |  | <0.001 |
| **Sex at birth (19,810)** |  |  |  |  |  |  |  |  |
| Man | 1,934 (98.6) | 16,113(90.3) | 1 |  |  | 1 |  |  |
| Woman | 27 (1.4) | 1,736 (9.7) | 0.15 | 0.1-0.2 | <0.001 | 0.3 | 0.2-0.5 | <0.001 |
| **Sexual orientation (19,820)** |  |  |  |  |  |  |  |  |
| Heterosexual | 76 (3.9) | 2,813 (15.7) | 1 |  |  | 1 |  |  |
| Homosexual/Gay/Lesbian | 1,722 (87.8) | 13,373(74.9) | 4.3 | 3.4-5.4 | <0.001 | 2.7 | 2.0-3.7 | <0.001 |
| Bisexual | 164 (8.3) | 1,672 (9.4) | 3.4 | 2.6-4.4 | <0.001 | 1.9 | 1.4-2.7 | <0.001 |
| **Gender identity (19,820)** |  |  |  |  |  |  |  |  |
| Cisgender | 1,889 (96.3) | 17,423(97.6) | 1 |  |  |  |  |  |
| Transgender | 60 (3.0) | 355 (2.0) | 1.5 | 1.2-1.8 | <0.001 |  |  |  |
| Travestis | 13 (0.7) | 80 (0.4) | 1.4 | 0.9-2.4 | 0.174 |  |  |  |
| **Ethnicity/skin color (19,816)** |  |  |  |  |  |  |  |  |
| Pardo (Brown, i.e., mixed race) | 650 (33.1) | 5,436 (30.4) | 1 |  |  | 1 |  |  |
| White | 1,043 (53.2) | 9,964 (55.8) | 0.9 | 0.8-0.98 | 0.02 | 0.8 | 0.7-0.9 | <0.001 |
| Black | 234 (11.9) | 2,192 (12.3) | 0.9 | 0.8-1.1 | 0.18 | 0.8 | 0.7-0.99 | 0.04 |
| Yellow (i.e., Asian Brazilian) | 25 (1.3) | 199 (1.1) | 1.04 | 0.8-1.5 | 0.83 | 1.0 | 0.6-1.5 | 0.97 |
| Indigenous | 9 (0.4) | 64 (0.3) | 1.15 | 0.5-2.1 | 0.67 | 1.2 | 0.6-2.4 | 0.56 |
| **Schooling level (19,817)** |  |  |  |  |  |  |  |  |
| 8-11 years | 419 (21.0) | 4,171 (23.4) | 1 |  |  | 1 |  |  |
| None/No formal education | 1 (<0.1) | 24 (0.1) | 0.4 | 0.1-3.0 | 0.374 |  |  |  |
| 1-3 years | 6 (0.3) | 77 (0.2) | 0.8 | 0.4-1.7 | 0.550 |  |  |  |
| 4-7 years | 46 (2.3) | 669 (3.7) | 0.7 | 0.5-0.9 | 0.017 |  |  |  |
| 12 years and over | 1,490 (76.0) | 12,914(72.3) | 1.1 | 1.0-1.6 | 0.017 |  |  |  |
| **Accepted something in exchange for sex in the last 6 months (19,819)** |  |  |  |  |  |  |  |  |
| Yes | 252 (13.0) | 1,484 (8.3) | 1.5 | 1.3-1.7 | <0.001 | 1.6 | 1.3-1.9 | <0.001 |
| No | 1,710 (87.0) | 16,373(91.7) | 1 |  |  | 1 |  |  |
| **History of syphilis in the 6 months before PrEP (19,820)** |  |  |  |  |  |  |  |  |
| Yes | 318 (16.2) | 1,275 (7.1) | 2.2 | 2.0-2.5 | <0.001 | 2.2 | 1.9-2.5 | <0.001 |
| No/Unknown | 1,644 (83.8) | 16,583(92.9) | 1 |  | 1 |  |  |  |
| **Condom use in the last 3 months <sup>‡</sup>(18,824)** |  |  |  |  |  |  |  |  |
| Never | 408 (21.5) | 4,240 (25.0) | 1.3 | 1.1-1.5 | <0.001 | 1.2 | 1.1-1.5 | 0.01 |
| Less than half of the time | 397 (20.9) | 2,453 (14.5) | 2.1 | 1.8-2.4 | <0.001 | 1.7 | 1.5-2.0 | <0.001 |
| Half of the time | 281 (14.8) | 1,830 (10.8) | 2.0 | 1.7-2.3 | <0.001 | 1.7 | 1.5-2.1 | <0.001 |
| More than half of the time | 489 (25.7) | 3,895 (23.0) | 1.7 | 1.4-1.9 | <0.001 | 1.4 | 1.2-1.6 | <0.001 |
| Always | 324 (17.0) | 4,507 (26.7) | 1 |  |  | 1 |  |  |
| **Sexual partners in the last 3 months<sup>¥</sup>** |  |  |  |  |  |  |  |  |
| Men | 18 ± 230 (4) | 8 ± 36 (2) |  |  | <0.001 |  |  | <0.001 |
| Women | 0.11±1.36(0) | 0.20 ± 2.45(0) |  |  | <0.001 |  |  | 0.29 |
| Trans women | 0±0.05(0) | 0.01±0.25 (0) |  |  | 0.058 |  |  | 0.19 |
| Trans men | 0.01±0.13(0) | 0.01±0.43(0) |  |  | 0.006 |  |  |  |
| Travestis | 0.01±0.35(0) | 0.01± 0.54(0) |  |  | 0.944 |  |  |  |
| **Alcohol use<sup>¥\$</sup>(18,979)** | | | | | | | | |
| Yes | 912 (47.0) | 7,367 (43.0) | 6.7 | 6.3-7.0 | <0.001 |  |  |  |
| No | 1,009 (53.0) | 9,691 (57.0) | 1 |  |  |  |  |  |
| **Use of injectable drugs<sup>¥</sup>(18,954)** |  |  |  |  |  |  |  |  |
| Yes | 11 (0.6) | 72 (0.4) | 1.3 | 0.7-2.3 | 0.348 |  |  |  |
| No | 1,913 (99.4) | 16,958 (99.6) | 1 |  |  |  |  |  |
| **Uso of other psychoactive drugs** |  |  |  |  |  |  |  |  |
| Poppers | 120 (6.1) | 425 (2.4) | 2.5 | 2.1-2.9 | <0.001 | 1.5 | 1.2-1.9 | <0.001 |
| Cocaine | 210 (11.0) | 905 (5.1) | 2.1 | 1.8-2.4 | <0.001 | 1.5 | 1.2-1.7 | <0.001 |
| Crack | 1 (<0.1) | 26 (0.1) | 0.4 | 0.1-2.8 | 0.597 |  |  |  |
| Marijuana | 425 (22.0) | 2,716 (15.0) | 1.5 | 1.4-1.7 | <0.001 | 1.2 | 1.01-1.3 | 0.03 |
| Club drugs (MDMA, GHB, ketamine, roofies, methamphetamine, and LSD) | 216 (11.0) | 1,237 (6.9) | 1.7 | 1.5-1.9 | <0.001 |  |  |  |
| Erection stimulants | 220 (11.0) | 1,126 (6.3) | 1.8 | 1.6-2.1 | <0.001 | 1.1 | 0.96-1.3 | 0.14 |
| Solvents | 22 (1.1) | 101 (0.6) | 2.0 | 1.4-3.0 | <0.001 |  |  |  |

\* RR: Relative Risk \*\* Mean ± Standard Deviation (Median); \*\*\* Chi-square test;
‡ Data from the last follow-up appointment § 5 doses or more in two hours; % Fischer exact test .# aOR: Adjusted Odds Ratio ## 95%CI: 95% Confidence Interval.
Akaike Information Criterion (AIC) = 11.367; pseudoR<sup>2</sup> = 0.157.

In the bivariate analysis, statistically significant differences were observed in the following variables: sex at birth, sexual orientation, gender identity (transgender), ethnicity/skin color, schooling years (4-7 years and 12 years or more), having accepted something in exchange for sex, history of previous syphilis in the last six months, frequency of condom use in the last three months, number of sexual partners in the last three months, and alcohol and other psychoactive drugs abuse in the last three months.

The multivariate analysis (see Table 3) identified no multicollinearity based on VIF (see Supplement 2). After we identified the most parsimonious model, the variables independently associated with the incidence of syphilis in the cohort were age, sex at birth, self-reported ethnicity/skin color, having accepted something in exchange for sex, history of previous syphilis in the last six months, frequency of condom use in the last three months, number of sexual partners in the last three months (men, women, and transsexuals) and use of poppers, cocaine, marijuana, and erection stimulants.

Regarding the risk of acquired syphilis during PrEP use, being a woman was a protective factor (aOR 0.3; 95% CI 0.2-0.5). White and Black people had an 80% lower risk of having syphilis than brown people (95% CI 0.7-0.9 and 0.7-0.99, respectively). Homosexuals, gays, and lesbians were 2.7 times (95% CI 2.0-3.7) more likely to be at risk of having syphilis than heterosexuals. Moreover, individuals with a history of syphilis in the six months before PrEP were 2.2 times (95% CI 1.9-2.5) more likely to be at risk of acquiring syphilis during follow-up.

Regarding behaviors reported at the last follow-up visit, people who accepted something in exchange for sex were 1.6 times (95% CI 1.3-1.9) more likely to be at risk of having syphilis than individuals who did not engage in this practice. Irregular condom use increased by at least 1.2 times the risk compared to those who use condoms in all relationships. The use of recreational drugs such as poppers (aOR 1.5; 95% CI 1.2-1.9), cocaine (aOR 1.5; 95% CI 1.2-1.7), and marijuana (aOR 1.2; 95% CI 1.01-1.3) is also associated with an increased risk of syphilis during PrEP use. There was also a difference between the groups regarding age and the number of male sexual partners.

## DISCUSSION

This research analyzed a cohort using digital records from different stages among PrEP users nationwide to understand the incidence of syphilis in the context of PrEP and its associated factors. Most PrEP users were male, white, self-identified as male, homosexual, gay or lesbian, and had 12 or more schooling years. Our findings revealed a prevalence of 8.3% of syphilis in the six months before starting PrEP, 4% in the first 30 days of prophylaxis use, and 9.8% of all those followed up in this cohort had syphilis during follow-up (with an incidence of 19.1 cases of acquired syphilis per 100 people), with 1.9% reinfected users.

The multivariate analysis identified female gender as a protective factor against syphilis during PrEP, as with being white or Black. On the other hand, the main risk factors for syphilis during PrEP were identified as being homosexual, gay, or lesbian and having a history of syphilis in the six months before starting PrEP. Risk behaviors associated with syphilis included accepting something in exchange for sex, irregular condom use, and recreational drug use. We also observed significant differences between the groups regarding age and the number of male sexual partners. Moreover, we noted a significant proportion of follow-up appointments without testing or with unavailable tests.

Our findings estimated an 8.3% prevalence of syphilis in the six months before starting PrEP. Recent studies of PrEP users in other countries have revealed rates of 10.1% (95% CI 7.6-13.1%) in Canada [13], 3.63% (95% CI 2.93-4.44%) in Germany [14] and 25.8% in France [15]. However, it is essential to exercise caution when comparing these studies since the approach to analyzing the history of syphilis before PrEP can vary (interview, history of laboratory tests, and medical records), which can directly influence the prevalence estimates found.

The follow-up of PrEP users in Brazil revealed an incidence of 19.1 cases of acquired syphilis per 100 person-years (95% CI 18.4-19.9). This incidence can be considered relatively high against results from other cohorts of PrEP users in Australia (9.4 (9.0-9.8) /100 PA) [16], Canada (1.94 (0.73-5.12) /100 BP) [13], and the United States (7.8 (7.1-8.4) /100 BP) [17], but similar to findings in Denmark (21.7 /100 BP) [18] and Spain (15.99 (11.94-21.41) /100PA) [19]. These regional disparities need to be better investigated. They may reflect vulnerabilities arising from the response of the countries’ public policies in the fight against syphilis and the profile of PrEP users in each country.

While our findings found 1.9% reinfection, Lemmet et al [15] showed 17.1% of PrEP users followed up in France with reinfection, mainly in older people (aOR 1.31; 95% CI 1.04-1.67) and MSM (aOR 2.71; 95% CI 1.89-3.96). However, syphilis reinfection in the context of PrEP is little explored in the studies. It is, therefore, recommended to explore the factors associated with syphilis reinfection in further studies. Our low rate can be explained by the low testing level detected during follow-up appointments, perhaps due to fewer referrals to primary care.

Regarding the factors associated with syphilis in the PrEP, the study by Lemmet et al [15] corroborates our findings, as it shows that female gender can be a protective factor, while people with syphilis in PrEP may be 6.6 more likely (CI 95% 5.10-8.66) to be men who have sex with men. Our findings also showed female gender as a protective factor, being homosexual, gay, or lesbian, and age as a risk factor. A study conducted by Peel et al [20] in Australia showed that PrEP users diagnosed with syphilis had a mean of 33 years and that, as in our findings, there is evidence that a syphilis history is a relevant risk factor [14, 15].

Regarding behaviors associated with syphilis infection during PrEP, our study found a significant association between recreational drug use and syphilis acquired during PrEP, particularly for poppers, marijuana, cocaine, and erection stimulants. One study identified that sexual drugs and chemsex have a significant association with the number of partners, search for partners on the internet, and low condom use with casual partners [21]. These findings may corroborate an increased vulnerability with the use of these substances. Hetcher et al [17] emphasizes that the relationship between the use of these substances and STIs needs to be examined more directly in order to understand the mechanisms that promote the vulnerability of individuals when they are using these substances. Indeed, understanding how these fragilities operate in the health-disease process for syphilis can facilitate management during PrEP to prevent this infection and other STIs.

Although the highest proportion of PrEP users are white (53.7%), this category of the ethnicity/skin color variable has an 80% lower risk of syphilis infection than brown people. In the Brazilian population, most people self-declare brown (45.3%) or white (43.5%) [22], and this can be investigated in future studies in light of the unequal access to PrEP in Brazil. In Brazil, equitable access by law to health resources through the health decentralization policy of the Unified Health System has significant challenges, considering that it offers a universal system to more than 180 million inhabitants. The unequal distribution of health resources is a persistent reality in several contexts and is a complex issue [23]. However, the possible unequal distribution of PrEP would occur not only in Brazil but also among Americans [24].

Furthermore, we observed significant follow-up appointments without testing or unavailable tests. The COVID-19 pandemic may have impacted the quality of care involving serological testing [25], which started in 2020 since the provision of services during this period was adversely affected, with outpatient sexual and reproductive health services adopting models of care that avoided face-to-face appointments and hospitals with shortened appointments in order to reduce the number of people in hospital environments. Moreover, people from the lesbian, gay, bisexual, transgender, queer, intersex, and agender (LGBTQIA+) community, a primary population for PrEP, can face barriers in health services due to misgendering, stigma, and attitudes that can generate bad experiences for them, interfering with their healthcare [26]. Individual service and social issues, such as improving the convenience of testing through self-testing and offering a service that men feel comfortable and safe accessing, should be considered by health services to make STI testing more accessible to MSM [27]. Serological testing for both syphilis and other STIs is a relatively simple and low-cost technology in the care of PrEP users. It should be prioritized in the context of inputs for primary health services.

Differentiated approaches to syphilis should be considered for this specific subgroup of PreP users. Initially, we could not observe in this Brazilian research an approach to communication with sexual partners for testing and treatment in order to interrupt the chain of transmission of syphilis, although recommendations are in place in this country and elsewhere in the world [9, 28]. Also, the use of social media apps for sexual encounters can favor anonymity and an increase in the number of partners [29]. However, this variable may not be addressed during follow-up appointments as it is not included in the guide form for monitoring appointments. It is, therefore, recommended to explore new approaches within PrEP to reduce STIs.

Our study has some limitations. Firstly, the proportion of losses in the cohort was high (41%). However, the individual characteristics of those lost to follow-up were similar, proportionally, to those who continued (homosexuals, whites, and with a high schooling level). Secondly, the behavioral variables during follow-up were extracted only from the last visit, which may not have captured changes during PrEP use and the possible influence of these changes on *Treponema pallidum* infection. We recommend studies that analyze the behavior of PrEP users over time. Thirdly, some data were not recorded at the last visit, which can introduce information bias, and a significant proportion of tests was not run on appointments, which can underestimate the actual incidence density of acquired syphilis. Finally, as the data were obtained through interviews conducted by health professionals, there may have been inhibition in responses and underestimation of behaviors due to social desirability. Given the users’ schooling level, we should consider using self-administered questionnaires to reduce this bias.

These limitations underscore the need for caution in interpreting the results and highlight areas for future research aimed at improving understanding of the factors associated with syphilis in PrEP users and improving the quality of the data collected during follow-up of these individuals. Despite the limitations, our study provides substantial evidence for understanding syphilis during HIV prophylaxis. Besides the high rates of acquired syphilis, the findings related to associated factors can contribute to developing specific public policies geared to the health of homosexual men, those with a history of syphilis, a high number of sexual partnerships, and recreational drug users, curbing syphilis transmission rates and achieving the 2030 Agenda global goals.

We should underscore that the high incidence of syphilis among PrEP users should not be seen as a disadvantage of this form of prophylaxis, which could lead to compensatory risk behaviors. This situation requires additional studies beyond secondary data to explore other associated factors. Thus, the detection of syphilis and other STIs should be an integral part of managing these users during follow-up, and PrEP is an essential policy for reducing STI rates.

## Data Availability

Data cannot be shared publicly due to the individualized nature of PrEP user data. However, the data are available upon request from the databases via email to the corresponding author, for researchers who meet the criteria for access to confidential data.

## Notes

### Competing Interest Statement

The authors have declared no competing interest.

### Author Declarations

Ethics Committee of the Faculty of Medicine of the University of Brasília (CAAE: 07448818.0.1001.5558) The consent was not obtained because the data were analyzed anonymously

